# Individualized risk trajectories for iron-related adverse outcomes in repeat blood donors

**DOI:** 10.1101/2021.10.09.21264792

**Authors:** W. Alton Russell, David Schienker, Brian Custer

## Abstract

**Background:** Despite a fingerstick hemoglobin requirement and 56-day minimum donation interval, repeat blood donation continues to cause and exacerbate iron deficiency.

**Study design and methods:** Using data from the REDS-II Donor Iron Status Evaluation study, we developed multiclass prediction models to estimate the competing risk of hemoglobin deferral and collecting blood from a donor with sufficient hemoglobin but low or absent underlying iron stores. We compared models developed with and without two biomarkers not routinely measured in most blood centers: ferritin and soluble transferrin receptor. We generated and analyzed ‘individual risk trajectories’: estimates of how each donors’ risk developed as a function of the time interval until their next donation attempt.

**Results:** With standard biomarkers, the top model had a multiclass area under the receiver operator characteristic curve (AUC) of 77.6% (95% CI 77.3% - 77.8%). With extra biomarkers, multiclass AUC increased to 82.8% (95% CI 82.5% - 83.1%). In the extra biomarkers model, ferritin was the single most important variable, followed by the donation interval. We identified three risk archetypes: ‘fast recoverers’ (<10% risk of any adverse outcome on post-donation day 56), ‘slow recoverers’ (>60% adverse outcome risk on day 56 that declines to <35% by day 250), and ‘chronic high-risk’ (>85% risk of adverse outcome on day 250).

**Discussion:** A longer donation interval reduced estimated risk of iron-related adverse events for most donors, but risk remained high for some. Tailoring safeguards to individual risk estimates could reduce blood collections from donors with low or absent iron stores.

## INTRODUCTION

Repeat blood donation can cause or exacerbate iron deficiency, with higher incidence among teen donors and premenopausal women [1–6]. In the United States, potential donors are screened using fingerstick hemoglobin or hematocrit tests and deferred if levels are below a minimum cutoff. Currently, minimum hemoglobin levels are 12.5 g/dL for women and 13.0 g/dL for men. Because fingerstick hemoglobin is an unreliable indicator of iron stores, some donors with low or absent iron stores qualify to donate and are subjected to further iron loss [4]. In addition, low hemoglobin deferrals consume time and resources for both donors and blood centers, decrease donor satisfaction, and reduce the likelihood of future donations [7]. More reliable measures of iron status include ferritin, zinc protoporphyrin, soluble transferrin receptor, and hepcidin, but these are more costly to measure, and most are not yet available as point of care tests [8].

Past studies have identified several factors that increase risk of iron deficiency among blood donors. The Danish Blood Donor Study found that sex, menopause status, and donation history were the strongest predictors of iron deficiency among donors, and weight, age, vitamin use, and diet were also significant [5]. Similar results have been found for donors in the United States, Australia, and the Netherlands [1–4,6]. Other studies identified age, time since last donation, and donation history as strong predictors of a low hemoglobin deferral for repeat blood donors [9,10]. To our knowledge, no prediction model has been developed that considers the competing risks of hemoglobin deferral and of collecting blood from a donor with sufficient hemoglobin but low or absent underlying iron stores.

In this study, we used data for a cohort of donors from the REDS-II Iron Status Evaluation (RISE) study [11] to develop machine learning models that estimate the risks of hemoglobin deferral and collecting blood from a donor with low or absent iron stores as a function of the donation interval – the length of time from an index donation until the donor returns for a subsequent donation attempt. We analyzed the models’ predictive performance and variable importance, and we used the models to generate and assess donors’ risk trajectories.

## METHODS

Using data from the RISE study, we trained multiclass prediction models to predict the risk of three iron-related adverse outcomes at a subsequent donation attempt: hemoglobin deferral, donating with low iron stores, and donating with absent iron stores. We assessed the models’ predictive performance, compared performance with and without the inclusion of two non-routine biomarkers (ferritin and STfR) as features for prediction, and generated and analyzed individual risk profiles for each donor’s likelihood of iron-related adverse donation outcomes at their next visit as a function of their donation interval (how long until the donor returns). We have shared all code in a public repository [12] and provide the TRIPOD checklist (**Table S4**) [13].

### Data preprocessing and formatting

The RISE dataset contains data from six U.S. blood centers on donation attempts for 2,425 donors over a 2-year period December 2007 – December 2009 [11]. Enrolled participants completed a whole blood donation and agreed to donate frequently over the next two years. The study targeted equal numbers of male and female donors and about twice as many frequent donors compared to first time or reactivated donors. Collected data elements include donation history, biometrics for each visit, and questionnaire responses regarding demographics, diet, supplemental iron consumption, female reproductive health, and demographics. For the ‘standard biomarkers’ model, we used 46 variables available for donations in the RISE dataset together with the time interval until the donor returns to predict the outcome of a follow-up donation attempt. We assumed that donor characteristics measured at the baseline visit such as diet, vitamin use, smoking, and female reproductive health indicators would not change significantly over the study period, and we used them to predict outcomes following subsequent donations by the same donor. We also developed an ‘extra biomarkers’ model, for which we included ferritin, STfRr, and derived measures (log ferritin, ratio of STfR to log ferritin, and calculated body iron) as features for prediction. We re-coded or imputed missing values for some fields; **Table S1** contains these details for all features used for prediction. We also included a composite dietary iron consumption score that was generated for each donor in the RISE dataset as part of a prior secondary analysis of this dataset [14].

To generate the model development dataset, we considered donations with at least 150 mL of red blood cell loss as potential index donations, which included whole blood donations, mixed apheresis donations that included a single red cell unit, and some donations that were classified as ‘quantity not sufficient.’ We excluded potential index donations that were double red cell donations due to limited data, the altered iron recovery profiles that follows the large iron loss from double red collection, and the 112-day mandatory deferral period after such donations. We also excluded donations that were missing a measurement of ferritin and donations for which neither fingerstick hemoglobin nor hematocrit was recorded. If follow-up visits were recorded after potential index donations, we generated labels with the time until the follow-up visit (in days) and its outcome. For all index donations followed by a visit with significant iron loss, defined as a loss of at least 55 mL of red blood cells, we generated a label for the index donation based on the first such follow-up visit. Additionally, we generated labels for any follow-up visits that did not result in significant iron loss (i.e., visits resulting in a deferral or apheresis donations of platelets or plasma with <55 mL of red blood cell loss) if such visits occurred between the index donation and the first follow-up visit with significant iron loss. For each index donation *i*, the outcome of its follow-up visits (*z*_*i*_) was classified as hemoglobin deferral (labeled as *z*_*i*_ = 1) if one were recorded; as a low iron donation (*z*_*i*_ = 2) if pre-donation ferritin was ≥ 12 mg/dl and < 20 mg/dl for women or ≥ 12 mg/dl and < 30 mg/dl for men; as an absent iron donation (*z*_*i*_ = 3) if pre-donation ferritin was <12 mg/dl; and as a ‘no adverse outcome’ donation otherwise (*z*_*i*_ = 0). Follow-up donations without ferritin measurements (*z*_*i*_ = −1) were not included in the model development dataset but were included in a ‘first return’ dataset. We used the first return dataset to calibrate the model and generate risk trajectories as described below.

### Prediction model development

#### Model selection

We evaluated several candidate model types: gradient boosted machines, random forest, regression trees, and generalized linear models with elastic net regularization with and without second order interaction terms. For each model type, to optimize performance while minimizing overfitting, we evaluated multiple parameter configurations via grid search with nested cross validation and resampling (**Table S2**) [15]. We generated 15 *model assessment partitions* which consisted of 3 resamples of 5 equal-sized partitions of the entire dataset, which we generated with stratified sampling to ensure the distribution of outcomes was balanced across partitions. For each model assessment partition, we defined all data not included in the partition as the corresponding *model tuning set*. Within the 15 tuning sets, we assessed all candidate model configurations (model type and hyperparameter setting) using 5-fold validation, assessing the multiclass area under the reliever operator characteristic curve (multiclass AUC) using the Hand and Till method [16]. We compared model configurations based on the average multiclass AUC across 5 cross validation folds averaged over all 15 tuning sets (assessing a total of 75 realizations of each candidate model configuration).

We also evaluated ensemble models, which combine the risk scores from multiple base models. We assessed two methods of combining risk scores from base models: a simple average and a weighted average, for which we weighted each model’s score proportionally to its accuracy raised to a power of four as suggested by Large et. al. [17]. We assessed AUC for each candidate ensemble configuration across the same 5 cross validation folds within each of the 15 tuning sets.

We selected the top model configuration based on multiclass AUC. To produce an unbiased assessment of the selected model configuration, we then assessed multiclass AUC on each of the 15 model assessment partitions. For each assessment partition, we trained the model configuration on all data not in the partition and used this model to generate risk scores on the assessment partition; we used those risk scores to calculate multiclass AUC. We completed this model development process both with ferritin, STfR, and derived measures as features (extra biomarkers model) and without (standard biomarkers model). We also computed one-vs-rest AUC for each feature, a measure of how well the model discriminates one outcomes from the other three.

#### Feature importance

For the top-performing “standard” and “extra biomarkers” model configurations, we assessed the importance of features for prediction using a random permutation method [18]. We trained the model on an altered version of each model tuning sets for which one feature column was randomly shuffled. We then generated risk scores for the corresponding model assessment partition and calculated the multiclass AUC. We calculated the percent decrease in multiclass AUC when a feature’s column was shuffled as compared to using the unaltered model tuning sets, which we used as a measure of the feature’s importance to the model.

#### Calibration

To generate the final model, we retrained the selected model configurations on the entire model development dataset, then we calibrated the predicted probabilities to the ‘first return’ dataset. In this dataset, index donations were labeled only once with the outcome of the first subsequent donation attempt, which included follow-up donations with no ferritin measurement. We estimated the distribution of outcomes in this dataset by assuming that followup donations with no ferritin measurement would have the same distribution of absent, low, and ‘no-adverse outcome’ donations as did the follow-up donations for which ferritin was measured. Mathematical details are provided in the supplemental methods.

### Risk trajectory analysis

For each index donation, we generated a risk trajectory using the calibrated ‘extra biomarkers’ model by predicting the likelihood of each outcome at the donor’s next donation attempt for each possible follow-up donation interval between 56 and 250 days. We generated graphical representations of individual donors’ risk trajectories showing how the estimated of each adverse outcome evolves depending on the number of days until the donor returns. To illustrate differences in risk trajectories, we created three recovery archetypes: ‘fast recoverers’ (<10% risk of any adverse outcome on post-donation day 56), ‘slow recoverers’ (>60% adverse outcome risk on day 56 that declines to <35% by day 250), and ‘chronic high-risk’ (>85% risk of adverse outcome on day 250). In a separate subgroup analysis, we compared the mean and 95% confidence interval for the estimated risk of each adverse outcome as a function of the donation interval for groups of donors stratified by selected parameters.

## RESULTS

### Data processing

In the RISE dataset, a total of 7817 donations from 1922 donors were followed by at least one follow-up visit. We excluded 520 index donations because hemoglobin was not recorded, and we excluded a further 18 index donations from the first return dataset because the first follow-up visit with significant iron loss was less than 56 days later. The first return dataset contained 7279 index donations labeled with the outcome of the first follow-up donation. That outcome was a hemoglobin deferral for 636 index donations; a low-iron donation for 754; an absent iron donation for 568; no adverse outcome for 1340; and a completed donation with unknown iron status for 3981. The model development dataset included 3529 unique index donations from 1543 donors. 3149 index donations were labeled with one follow-up donation, 289 were labeled twice, and 91 were labeled with 3 or more follow-up visit outcomes (maximum of 8).

### Prediction model

Separately for the standard and extra biomarker versions, we evaluated 2,006 non-ensembled model configurations (model type and hyperparameter setting) and four enemble models. For both versions, the top-performing non-ensembled model was a gradient boosted machine (**Figure S1, Table S2**). The top-performing standard biomarkers model configuration was an ensemble model that averaged the risk scores for three gradient boosted machine and three random forest models; the top extra biomarkers configuration was an ensemble model that averaged risk scores for two gradient boosted machines, a random forest model, and two penalized regression models, one with second order interaction terms. Multiclass AUC for the top ensemble models assessed on the model assessment partitions was 77.6% (95% CI 77.3% - 77.8%) for the standard biomarkers model and 82.8% (95% CI 82.5% - 83.1%) for the extra biomarkers model (**Table 1**). For both the standard and extra biomarkers model, the top ensemble model had a higher mean AUC with lower standard error than each of the base models that comprised it across the model tuning sets (**Figure S2**). Both models had the highest discriminative performance for predicting no adverse outcome donations and the lowest discriminative performance for predicting low iron donations (**Figure 1**). Inclusion of the extra biomarkers had the greatest improvement in distinguishing low and absent iron donations from the other outcomes (one-vs-rest AUC increased 6.9% for low iron donations and 8.9% for absent iron donations; **Table 1**).

**Table 1:** Multiclass and one verses rest AUC by outcome for the top “standard” and “extra biomarker” model configuration as assessed on the model assessment partitions.

| AUC metric |  | Standard biomarkers model | Extra biomarkers model | Difference |
| --- | --- | --- | --- | --- |
| Multiclass AUC |  | 77.6% (77.3% - 77.8%) | 82.8% (82.5% - 83.1%) | 5.2% |
| One-vs-rest AUC | No adverse outcome | 86.6% (86.2% - 87.0%) | 91.1% (90.9% - 91.4%) | 4.6% |
|  | Hemoglobin deferral | 81.1% (80.6% - 81.5%) | 81.7% (81.3% - 82.1%) | 0.6% |
|  | Low iron donation | 72.6% (72.0% - 73.1%) | 79.5% (79.0% - 80.0%) | 6.9% |
|  | Absent iron donation | 76.9% (76.5% - 77.3%) | 85.8% (85.4% - 86.2%) | 8.9% |

**Figure 1:**
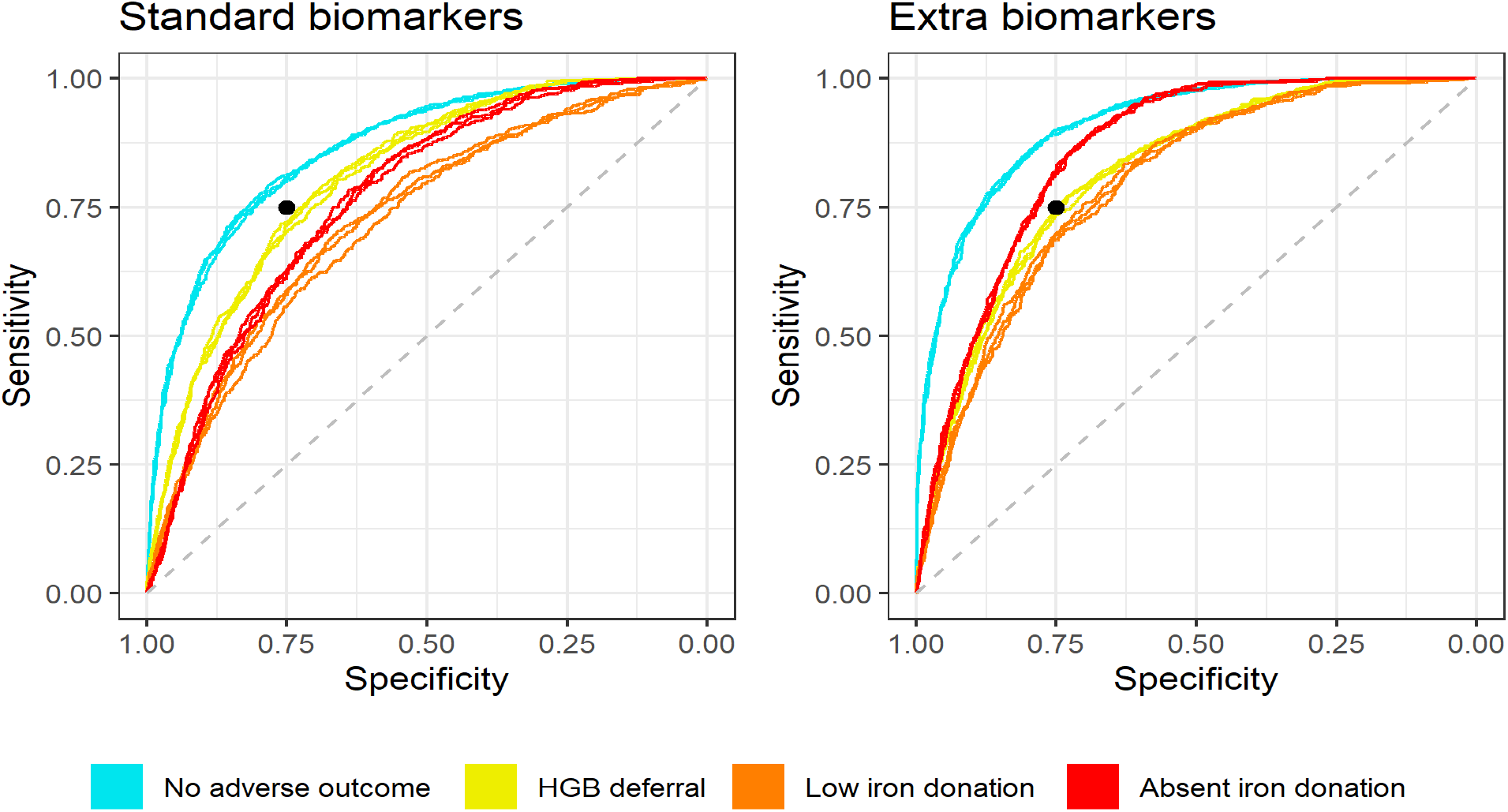
One-vs-rest ROC curves for the standard and extra biomarker models as assessed on the model assessment partitions. For each outcome, one ROC curve is plotted for each of the three resamples of the data, combining data from the corresponding 5 model assessment partitions. Black dot at 75% sensitivity and 75% specificity.

For the standard biomarkers model, the donation interval (time to return) was the most important feature for prediction (the median decrease in multiclass AUC when shuffling this feature was 4.9%), followed by venous hemoglobin and the number of red blood cell units donated in the last 24 months (median decreases in multiclass AUC of 3.1%, and 2.0%, respectively; **Figure 2** and **Figure S3**). For the extra biomarkers model, ferritin was by far the most important feature, followed by donation interval and fingerstick hemoglobin/hematocrit (median decreases in multiclass AUC of 3.6%, 1.5%, and 0.4%, respectively; **Figure 2** and **Figure S4**). For both versions, the calibration weights down-weighted relative likelihood of hemoglobin deferrals (**Table S*3***).

**Figure 2:**
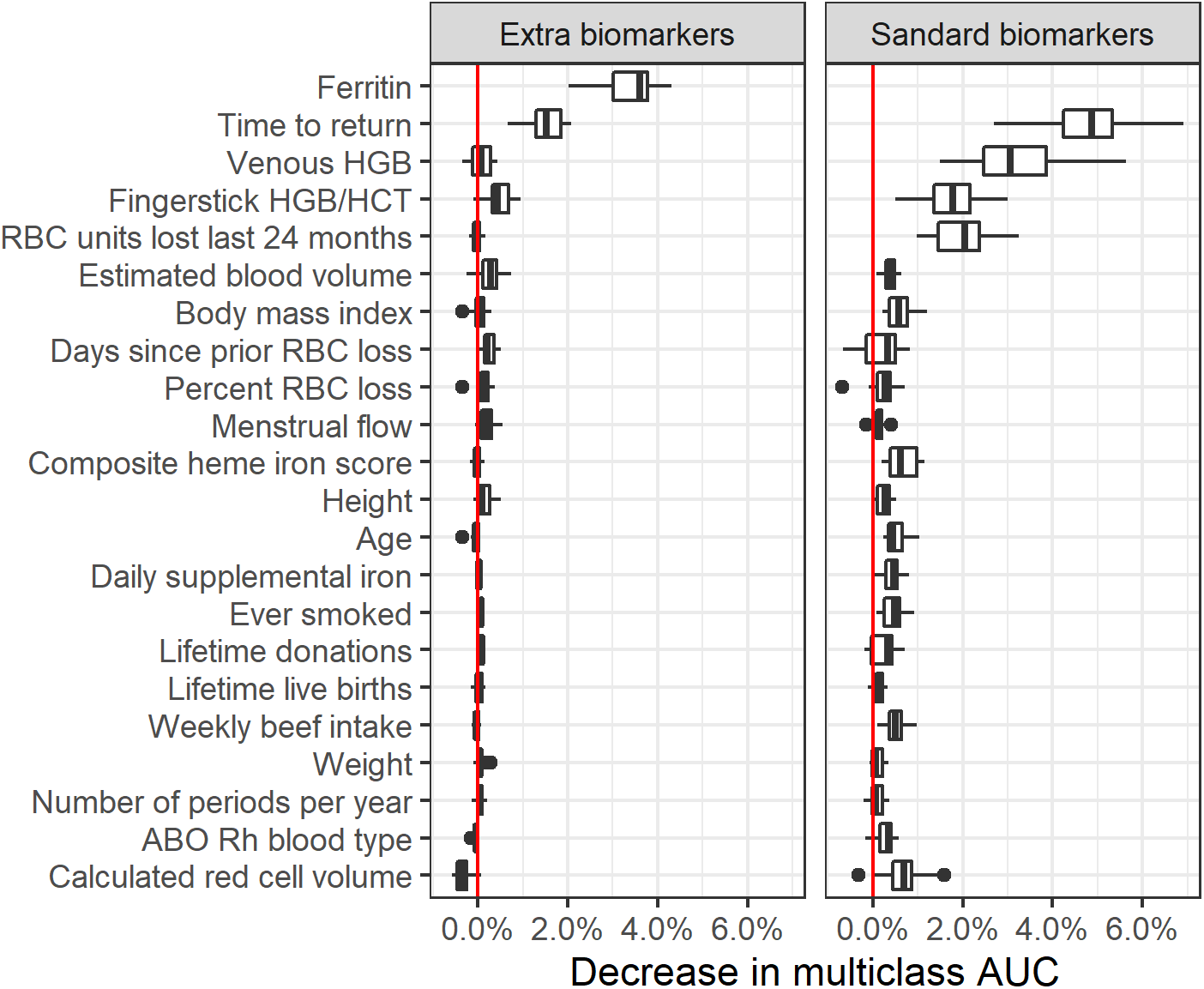
Relative variable importance for the top “standard” and “extra” biomarker models. Variables were included in this figure if among the top 15 most important variables for at least one of the models. Full variable importance plots shown in the supplement.

### Individual risk profiles

Using the calibrated extra biomarkers model on the first return dataset, the median risk of any adverse outcome with a 56-day donation interval was 69% (Interquartile range [IQR] 35% – 92%). For a 250-day interval, the median risk of any adverse outcome fell to 36% (IQR 11% – 66%). The median decrease in absolute risk from an interval of 56 to 250 days was 22% (IQR 9% – 32%).

Individual donor risk trajectories were highly heterogeneous (**Figure S5**). Across the first return dataset, 433 (12%) index donations were by a fast recoverer (<10% risk of any adverse outcome on post-donation day 56), 304 (8%) index donations were by a slow recoverer (>60% adverse outcome risk on day 56 and <35% on day 250), and 403 (11%) index donations were by a chronic high-risk donor (>85% risk of adverse outcome on day 250). Risk trajectories differed markedly across these three archetypes (**Figure 3**). For chronic high-risk donors, while overall adverse outcome risk slightly declined for longer donation intervals, risk of a low iron donation increased with donation interval for most donors (**Figure 4**).

**Figure 3:**
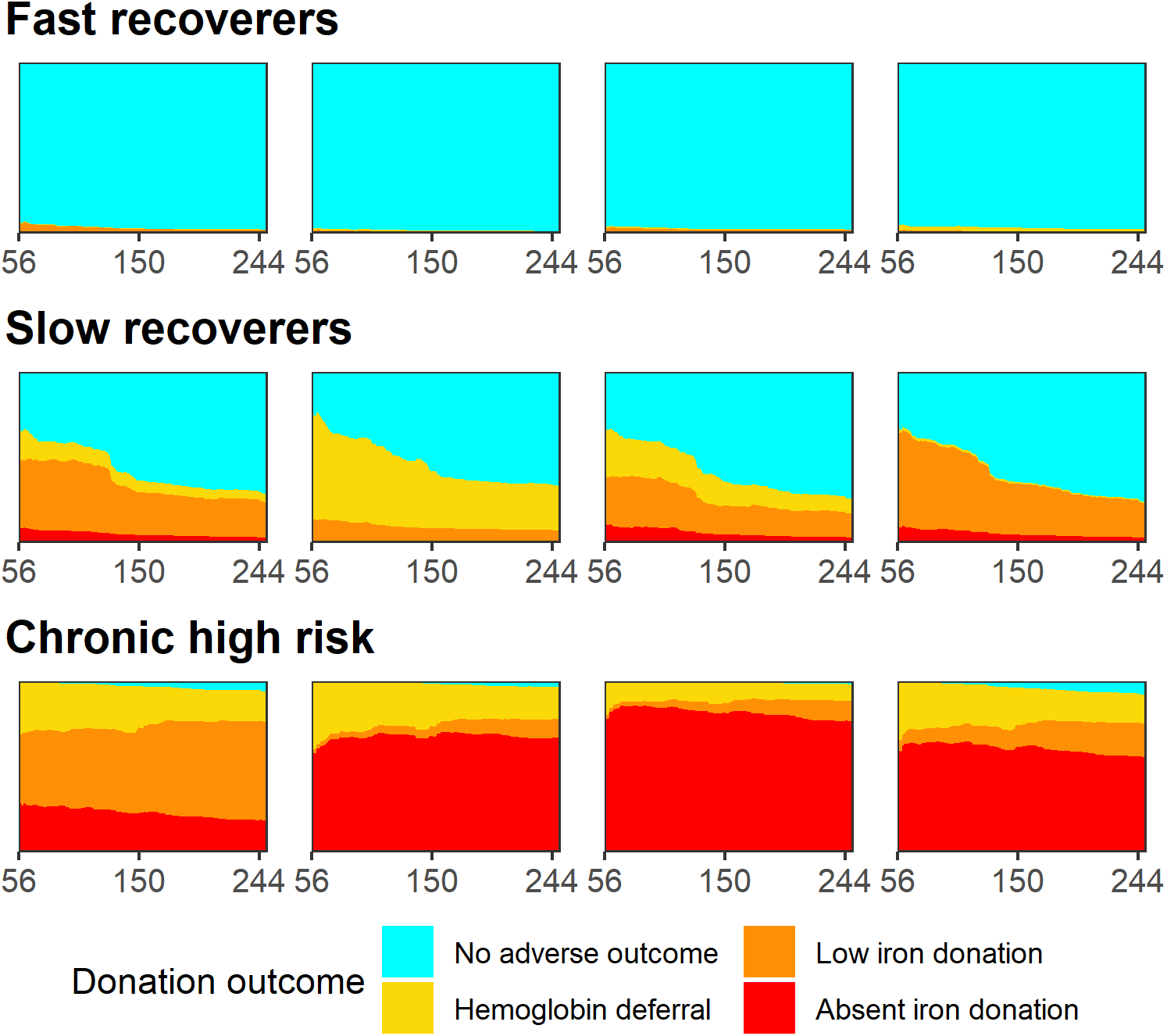
Individual risk profiles for four selected donors that represent each of the three donation archetypes. The donation interval (time to return donation attempt) is varied on the x axis from 56 to 250 days. Height of colored area indicates the risk of each adverse outcome and likelihood of a ‘no adverse outcome’ donation.

**Figure 4:**
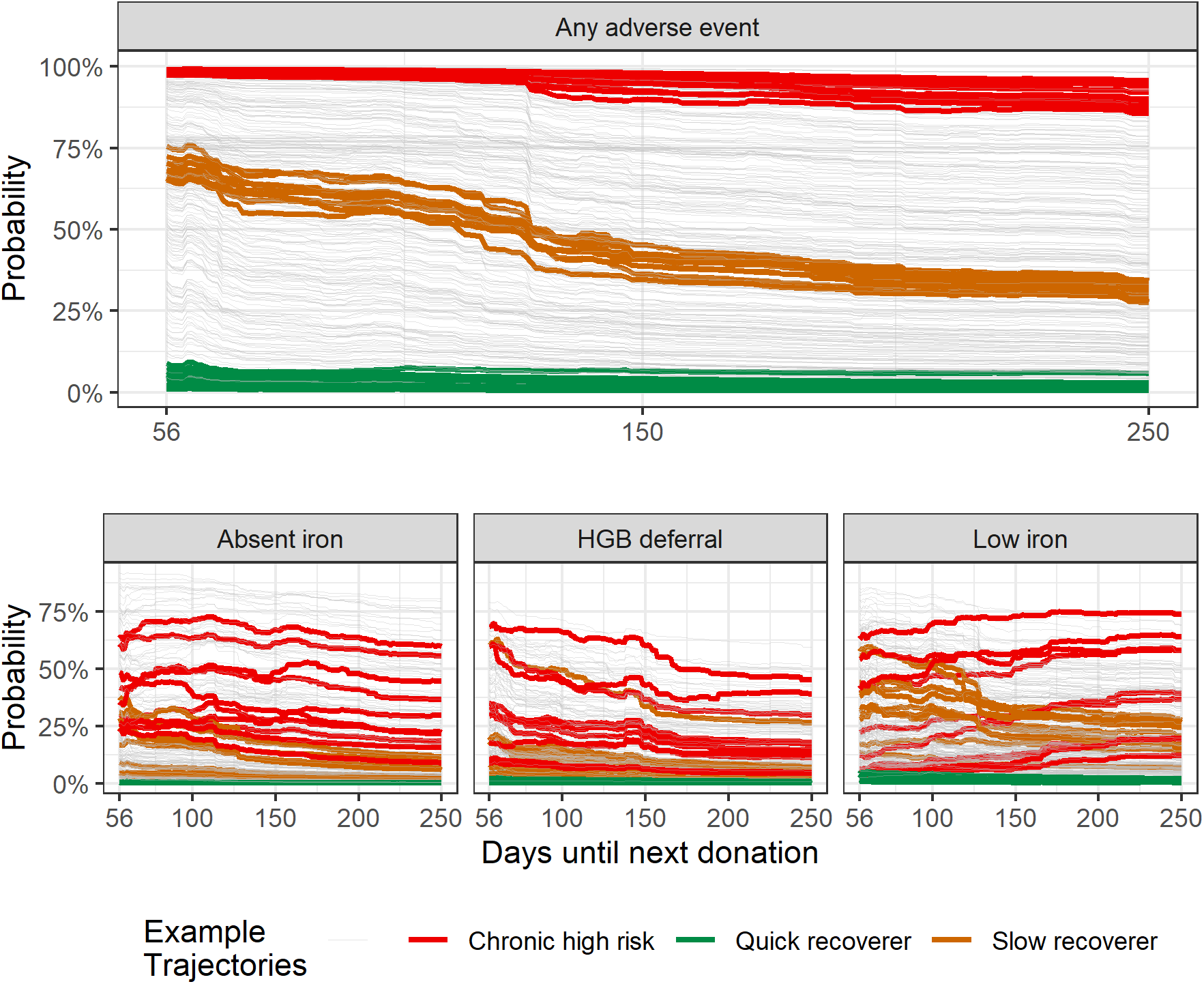
Risk trajectory for any adverse outcome (top plot) or a specific adverse outcome (bottom three plots) for 300 randomly selected donors. Five randomly selected donors fitting each of the three archetypes are highlighted in red, orange, and green. Other donors’ trajectories are shown in grey.

In cohort analysis, average risk of an adverse outcome was lowest for iron replete donors and highest for donors with absent iron at the index donation; average risk for low-iron donors was in between that of the other two cohorts (**Figure 5**). While overall adverse outcome risk declined with longer donation intervals for all three cohorts, risk of a low iron donation increased with longer intervals for donors with absent iron stores at the index donation. when defining cohorts based on the tertile of venous hemoglobin at the index donation, donors with venous hemoglobin in the lowest tertile (9.8-13 g/dL) had the highest risk of any adverse outcome (**Figure 6**). Whereas an absent iron donation was the most likely adverse outcome for a donor with absent iron stores at index donation, hemoglobin deferral was the most likely adverse outcome for a donor in the lowest tertile of venous hemoglobin at index donation. Average risk trajectory also differed across cohorts defined by gender (**Figure S6**), number of red blood cell units donated over the prior two years (**Figure S7**), self-reported iron supplementation use (**Figure S8**), and composite dietary heme iron intake (**Figure S9**).

**Figure 5:**
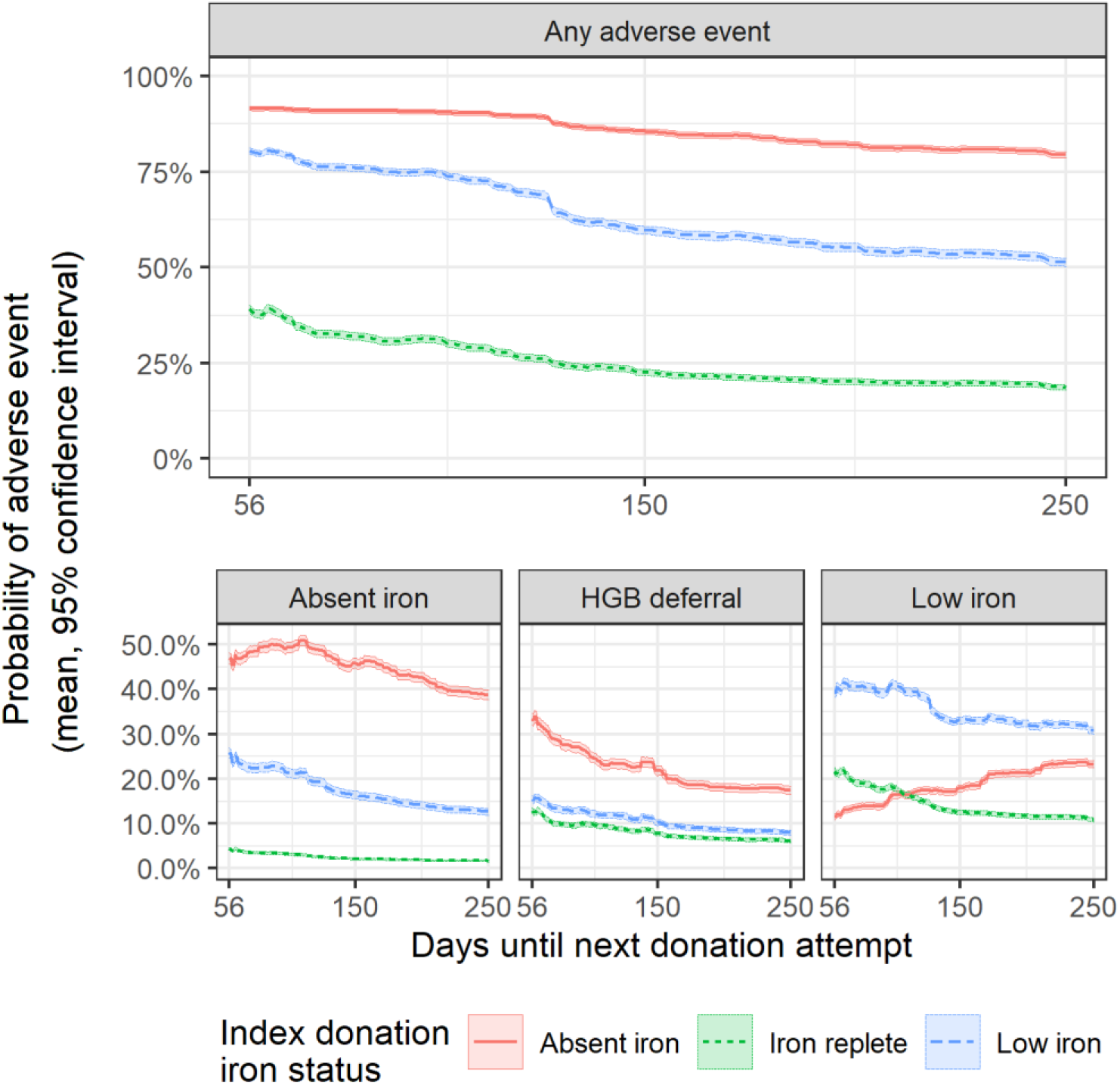
Average risk trajectory with 95% confidence intervals for donors in the first return dataset stratified by iron status at index donation, defined by the donor’s ferritin level.

**Figure 6:**
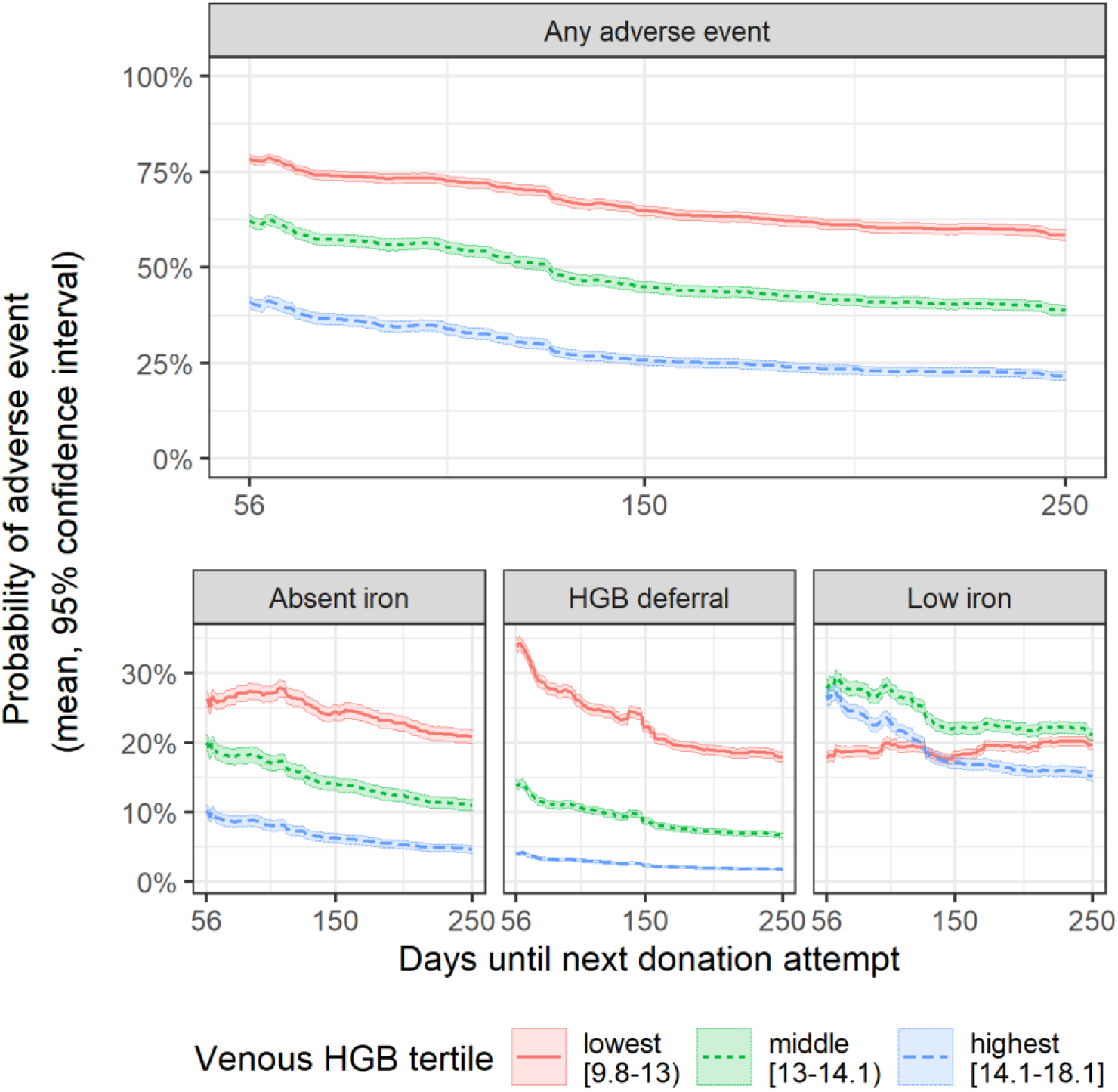
Average risk trajectory with 95% confidence intervals for donors in the first return dataset stratified by venous hemoglobin (HGB) measured at the index donation in g/dL.

## DISCUSSION

This analysis of 7279 index donations from the RISE study found that risk of iron-related adverse outcomes at follow-up donations can be estimated as a function of the interval before a follow-up donation attempt and that individual donors’ risk trajectories are highly heterogeneous. For most donors, estimated risk decreased precipitously if they waited longer to return, suggesting that longer minimum donation intervals would prevent some cases of donor-associated iron deficiency and hemoglobin deferrals. For some donors, risk remained high even with a 250-day donation interval. This heterogeneity in estimated risk trajectories suggests that uniform or sex-based intervals may be insufficient. Including ferritin as a predictor improved risk estimation, particularly with respect to estimating risk of absent iron donations. For some donors, estimated risk of an adverse outcome remained over 90% even for a 250-day donation interval. These donors may have underlying (and potentially undiagnosed) iron deficiency or a related condition, which may make them poor candidates for repeat blood donation. The heterogeneity in donor risk and the predictive power associated with the use of donor characteristics should be examined further in order to facilitate policy design such as personalized inter-donation intervals.

Our analysis has several limitations. Most notably, the RISE study population is not representative of a typical repeat blood donor population. RISE participants were asked to commit to frequent blood donation, and recruitment was targeted to achieve proportional representation based on sex and donation history [19]. We restricted our analysis to the subset of donations in the RISE study for which ferritin was measured, which may further bias our findings. Further study is needed to assess the generalizability of our prediction model’s performance to a more representative blood donor population. Many of the features we used for prediction are highly correlated (e.g., venous and fingerstick hemoglobin; 12- and 24-month donation history), which can cause feature importance to ‘spread’ over correlated features [20]. Due to this, our feature importance method should only be interpreted as which features the model relied on most (or was most sensitive to) rather than which features are most correlated with adverse outcome risk. To calibrate our model, we assumed the distribution of absent, low, and replete iron status for follow-up donations without a ferritin measurement mirrored the distribution across follow-up donations at which ferritin was measured, but this may not be the case.

We see several ways the approaches reported here can be used to gain further insights into tailored donation intervals for blood donors. Extension of this work to larger blood center operational datasets outside of specific clinical studies will provide information on the effectiveness of machine learning models when the quality and completeness of information may be more limited. Key features identified in this analysis are readily available such as donor hemoglobin/hematocrit, donation interval, and increasingly ferritin measurement. Other features such as venous hemoglobin and survey assessments of donor dietary habits and supplementation are not likely to be implemented as standard donor assessments.

Despite the limitations, our analysis demonstrates that repeat donors have heterogeneous risk of iron-related adverse outcomes as a function of their donation interval, and machine learning models can estimate individual donors’ risk trajectories. Such predictive models could be a valuable tool for managing risks to donors while ensuring a sufficient blood supply.

## Data Availability

The RISE dataset was accessed through the National Heart, Lung, and Blood Institute (NHLBI) Biolincc repository (https://biolincc.nhlbi.nih.gov). Our Research Materials Distribution Agreement prohibits publication of the raw data, but other researchers can submit a data request to NHLBI at no charge.

https://www.doi.org/10.5281/ZENODO.5247221

## DECLARATIONS

## Acknowledgments

The authors thank the NHLBI Biolincc repository for making the RISE dataset available at no charge, and we thank Dr. Bryan Spencer for providing the dietary heme iron intake scores he generated for a separate analysis.

## Ethics/Consent

Because our analysis used fully de-identified human subjects data for a secondary analysis, this study was exempted from full IRB review by the Stanford University IRB.

## Code availability

All code is uploaded to a public repository at https://www.doi.org/10.5281/ZENODO.5247221

## Authors’ contributions

All authors contributed to study design. WAR conducted the analysis and composed the manuscript; BC and DS edited the manuscript.

## SUPPLEMENTAL MATERIALS

### SUPPLEMENTAL METHODS

#### Calibration

Our calibration procedure was as follows: we totaled each follow-up outcome in the first return dataset as *n*^(*k*)^, where *k* = −1,0,1,2,3 correspond to a donation with unknown iron status (no ferritin measurement); a no adverse outcome donation, a hemoglobin deferral, a low iron donation, and an absent iron donation, respectively. We then calculated *ñ*^(*k*)^, an estimation of what the totals would have been if ferritin were measured for all follow-up donations assuming the distribution of outcomes was the same as for completed donations with ferritin measures. These were calculated as *ñ*^(1)^ = *n*^(1)^ (hemoglobin deferral) and 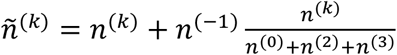 for *k* = 0,2,3 (completed donations). We then used our top model configuration to generate the unnormalized probability vector 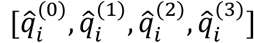 for each index donation *i* in the first return dataset. We computed weights *W*^(*k*)^ for the unnormalized probability of each outcome 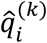 by solving the system of equations 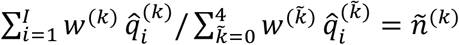 for each index donation *i ∈* 1,2, …, *l* and *k* = 0,1,2,3. The final calibrated model used parameters *w*^(*k*)^ together with the uncalibrated scores from the model 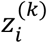 to produce the estimated likelihood of each outcome at a follow-up donation as 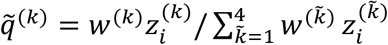 This ensured that the expectation of the distribution of the predicted outcome for the first return dataset would correspond to our estimated totals *ñ*^(*k*)^

### SUPPLEMENTAL TABLES

**Table S1:** List of features for prediction model with description and notes from feature engineering.

| Variable name | Description | Feature engineering |
| --- | --- | --- |
| **Donation history** |  |  |
| DER_RBC_Last12months | Red blood cell loss in last 12 months | Used as-is. |
| DER_RBC_Last24months | Red blood cell loss in last 24 months | Used as-is. |
| DER_RBCLoss_Units | Units of red blood cell lost at this donation | Used as-is. |
| DER_RBCLoss_mL | Volume of red blood cell loss at this donation (mL) | Used as-is. |
| DER_DaysRBCLoss | Days since last red blood cell loss | Set NAs (36%) to 10 years ago (10*365 days) |
| DER_DaysDRLoss | Days since last double red blood cell donation | Set NAs (36%) to 10 years ago (10*365 days) |
| RQ1_Ever_Donated | Ever donated blood before | Recoded to 0 (no), 1 (yes); set as 1 for all followup donations since they gave an index donation |
| cumLifetimeDonations | Total lifetime donations | If lifetime donations missing from baseline questionnaire, set to 0. For follow-up visits, increased by 1 for all donations. |
| **Biometric** |  |  |
| FingerstickHGB_equiv | Fingerstick HGB or HCT | Used fingerstick HGB if measured. Otherwise, used fingerstick HCT/3.04. If also missing, used adjusted venous HGB. |
| DD_ABO_RH | ABO-Rh blood type | 1 donor has UNT. Setting to most common value in dataset, O+ (40% of donors at baseline) |
| DER_AdjVenousHgb | Venous HGB (converted to pre-donation if measured from a post-donation sample) | Not collected at all follow-up visits. If missing, set to fingerstick value |
| DER_Weight | Weight (pounds) | Missing for 5 donors with 22 follow-up donations. 4 M, 1 F, ages 27 - 69. Imputing mean weight by gender among those aged >25 |
| DER_Height | Height (inches) | Missing for 6 donors (same 5 missing weight plus one additional) with 23 follow-up donations. Age 27 - 69. Imputing mean height by gender among those age >25. |
| BMI | Body Mass Index | Missing for same 6 donors missing height. Calculating from (imputed) weight and height |
| DER_EBV | Estimated Blood Volume (Nadler's equations) | None missing |
| DER_RedCellVolume | Total body red blood cell volume | Calculated total body red cell volume; none missing |
| DER_PercentRBCLoss | Estimated percent red blood cell volume loss at visit | Percent of red cell lost at donation; none missing |
| DER_Age | Age | Available for all, 18 - 87 |
| **Survey responses from baseline visit** |  |  |
| DD_Country | US born | 1 if US-born, 0 otherwise. Missing for <1%; if missing re-coding as US-born (>95%) |
| DD_Gender | Gender | no missing. 52% female at baseline; 51% for followups |
| DD_Raceth | Race/ethnicity (Asian, Black, Hispanic, Other, White) | 0.7% missing; Recoding to "O" (other) |
| RQ7_Ever_Smoked | Smoked at least 100 cigarettes in lifetime | Recoding to 0 (no) and 1 (yes). For "don't know" (26) and "missing" (10), coding to "no" because majority (60%) |
| RQ8_Smoked_Past_90Days | Smoked in last 90 days | Recoding to 0 no and 1 yes. If NA (60%) or don't know (3%), recoding to no (68% of respondents) |
| RQ11_Liver | Liver consumption (times per week) | Recoding as times per week 0 (never), 0.5 (<1/wk), 1 (1/wk), 2 (2/wk), 3.5 (3-4x/wk), 5.5 (5-6x/wk), 7 (daily), 14 (2x or more/day); 59 are 99 (blank/don't know); setting them to most common which was never |
| RQ11_Beef | Beef consumption (times per week) | Recoding as times per week 0 (never), 0.5 (<1/wk), 1 (1/wk), 2 (2/wk), 3.5 (3-4x/wk), 5.5 (5-6x/wk), 7 (daily), 14 (2x or more/day); 98 are 99 (blank/don't know); setting them to most common which was 2x/wk |
| RQ11_LPCT | Lamb, pork, chicken, or turkey consumption (times per week) | Recoding as times per week 0 (never), 0.5 (<1/wk), 1 (1/wk), 2 (2/wk), 3.5 (3-4x/wk), 5.5 (5-6x/wk), 7 (daily), 14 (2x or more/day); 59 are 99 (blank/don't know); setting them to most common which was 3.5x/wk |
| RQ11_Clams | Clams consumption (times per week) | Recoding as times per week 0 (never), 0.5 (<1/wk), 1 (1/wk), 2 (2/wk), 3.5 (3-4x/wk), 5.5 (5-6x/wk), 7 (daily), 14 (2x or more/day); 121 are 99 (blank/don't know); setting them to most common which was never |
| RQ11_OMSS | Oysters, mussels, shrimp, or sardines consumption (times per week) | Oysters, mussels, shrimp, sardines. Recoding as times per week 0 (never), 0.5 (<1/wk), 1 (1/wk), 2 (2/wk), 3.5 (3-4x/wk), 5.5 (5-6x/wk), 7 (daily), 14 (2x or more/day); 100 are 99 (blank/don't know); setting them to most common which was less than 1/wk |
| RQ11_OtrFish | Other fish consumption (times per week) | Recoding as times per week 0 (never), 0.5 (<1/wk), 1 (1/wk), 2 (2/wk), 3.5 (3-4x/wk), 5.5 (5-6x/wk), 7 (daily), 14 (2x or more/day); 38 are 99 (blank/don't know); setting them to most common which was <1x/wk |
| RQ11_Eggs | Egg consumption (times per week) | Recoding as times per week 0 (never), 0.5 (<1/wk), 1 (1/wk), 2 (2/wk), 3.5 (3-4x/wk), 5.5 (5-6x/wk), 7 (daily), 14 (2x or more/day); 62 are 99 (blank/don't know); setting them to most common which was 1x/wk |
| RQ11_Dairy | Dairy consumption (times per week) | Recoding as times per week 0 (never), 0.5 (<1/wk), 1 (1/wk), 2 (2/wk), 3.5 (3-4x/wk), 5.5 (5-6x/wk), 7 (daily), 14 (2x or more/day); 33 are 99 (blank/don't know); setting them to most common which was 1x/day |
| compositeIronScore | Composite dietary iron score | Missing for 13 at baseline. Imputing mean. |
| supp_iron_pct_of_daily | Supplemental iron | Value between 0 (no iron suppl) and 1 (daily iron suppl), where multivitamins or iron supplementation are both counted as supplemental iron. Computed as $\min(1, (\text{ironsupp\_per\_week} + \text{RQ12C\_MV\_WithIron\_YN} * \text{multivitamins\_per\_week}) / 7)$ |
| multivitamins_per_week | Multivitamin consumption (times per week) | Based on RQ12A_MultiVitamins_YN and RQ12B_MultiVitamins_How_Often. Recoding to times per week. "1: everyday" becomes 7; "2: 4-6x per week" becomes 5; "3: 1-3 days per week" becomes 2. Don't know/missing (51) set to daily (7/wk). If they answered yes to taking multivitamins but missing or don't know for how often, coding as 4-6x per week. |
| RQ17_NumberOfPeriods | Periods in last year | Original is NA for all men and all women who say their period stopped, and for one F 19yo donor who answered NA to menstrual status question, likely because she is pre-menstrual and that wasn't an option. Coding all NAs as 0. Also, equal to 99 (don't know) for 13 donors who said yes to having periods. Coding to the most common value which is 12. |
| RQ18_Menstrual_Flow | Menstrual flow intensity | Recoding as 0 for no period, 1 for spotting, 2 for very light, 3 for light, 4 for moderate, 5 for heavy, 6 for very heavy/gushing. 3 women who have periods had a '9', presumably don't know or refused to answer. Recoding to most common answer, which was 4 (moderate), |
| menstrual_flow_times_freq | Menstrual frequency and flow | $RQ19\_MenstrualFlow/5 * RQ17\_NumberOf\_Periods/12$ |
| RQ19_Ever_Pregnant | Ever pregnant | Originally NA for all men and 10 women. Recoding as 0 (no) and 1 (yes), all NAs coded as 0. |
| RQ20_NumberOfPregnancies | Number of pregnancies | Originally NA for all men and 356 women (same as never pregnant), 1-6 for other women. Coding NAs as 0. |
| RQ21_NumberOfLiveBirths | Number of live births | Originally NA for all men and 356 women; Coding NAs as 0. |
| gender_menstrating_cohorts | Gender & menstration cohort | 3 categories: male, female menstrating (>0 periods in last year), female not menstrating (0 periods in last year) |
| **Additional biometrics available for some donations** |  |  |
| ARUP_Ferritin | Ferritin (mg/dL) | Use as-is for extra biomarkers model |
| ARUP_STR | Soluble Transferrin Receptor (STfR) (nmol/L) | Use as-is for extra biomarkers model |
| DER_ARUP_log_Ferr | Log of ferritin | Use as-is for extra biomarkers model |
| DER_ARUP_log_STfR_Ferr | STfR divided by the log of ferritin | Use as-is for extra biomarkers model |
| DER_BodyIron | Body iron in mg/kg calculated from STfR | Use as-is for extra biomarkers model |

**Table S2:** Model types and hyperparameters assessed as candidates. All hyperparameter combinations were assessed in 5-fold cross validation on each of 15 model validation sets defined by the nested cross validation scheme. SB = Standard biomarkers version, XB = extra biomarkers version

| Hyperparameters | Values assessed | Top configuration hyperparameter value | Top configuration AUC (mean and range) |
| --- | --- | --- | --- |
| Gradient boosted decision trees (R package xgboost) |  |  |  |
| Learning rate | 0.01, 0.05, 0.1, 0.3 | SB: 0.01, XB: 0.05 | SB: 75.93% (75.41% - 76.45%)<br>XB: 82.05% (81.69% - 82.41%) |
| Maximum tree depth | 2, 4, 6, 8, 10, 12, 14, 16, 18, 20 | SB: 16, XB: 6 |  |
| Minimum child weight | 0, 1, 2, 4, 8 | SB: 2, XB: 8 |  |
| Row subsampling per tree | 0.65, 0.8, 1 | SB: 0.65, XB: 0.65 |  |
| column subsampling per tree | 0.8, 0.9, 1 | SB: 0.8, XB: 0.9 |  |
| Random forest (R package randomForest) |  |  |  |
| Minimum size of terminal nodes | 1, 2, 4, 8 | SB: 4, XB: 8 | SB: 75.90% (75.28% - 76.52%)<br>XB: 80.92% (80.52% - 81.33%) |
| Maximum number of trees | 250, 500, 750, 1000, 1250, 1500, 1750, 2000, 2250, 2500 | SB: 2500, XB: 2250 |  |
| Sampling with replacement? | yes, no | SB: yes, XB: no |  |
| Elasticnet penalized logistic regression (R package glmnet) |  |  |  |
| alpha (0 = ridge, 1 = lasso) | 0, .25, .5, .75, 1 | SB: 0, XB: 0 | SB: 72.49% (71.99% - 72.99%)<br>XB: 80.28% (79.85% - 80.71%) |
| lambda (penalty weight) | 0, 0.01, 0.02, 0.03, 0.04, 0.05, 0.06, 0.07, 0.08, 0.09, 0.1 | SB: 0.01, XB: 0 |  |
| Elasticnet with all second order interactions (R package glmnet) |  |  |  |
| alpha (0 = ridge, 1 = lasso) | 0, .25, .5, .75, 1 | SB: 0, XB: 0.75 | SB: 74.74% (74.22% - 75.26%)<br>XB: 82.05% (81.69% - 82.41%) |
| lambda (penalty weight) | 0, 0.01, 0.02, 0.03, 0.04, 0.05, 0.06, 0.07, 0.08, 0.09, 0.1 | SB: 0.04, XB: 0.01 |  |
| Regression trees (R package rpart) |  |  |  |
| Complexity parameter | 0.001, 0.005, 0.01, 0.05, 0.1 | SB: 0.001, XB: 0.001 | SB: 66.99% (66.32% - 67.65%)<br>XB: 75.59% (74.98% - 76.20%) |
| Minimum observations per split | 10, 15, 20, 25, 30 | SB: 30, XB: 30 |  |

**Table S3:** Calibration weights calculated for matching the expected distribution of the risk scores to the estimated distribution in the first return dataset. Compared to the raw risk predictions generated by the model trained in the model development dataset, calibration down-weighted risk of hemoglobin deferral (evidenced by a calibration weight less than 1) and up-weighted likelihood of the other three outcomes for both models.

| Model version | No adverse outcome | Hemoglobin deferral | Low iron donation | Absent iron donation |
| --- | --- | --- | --- | --- |
| Standard biomarkers | 1.044957 | 0.7297557 | 1.218854 | 1.190322 |
| Extra biomarkers | 1.046844 | 0.7358226 | 1.128668 | 1.204911 |

**Table S4:**
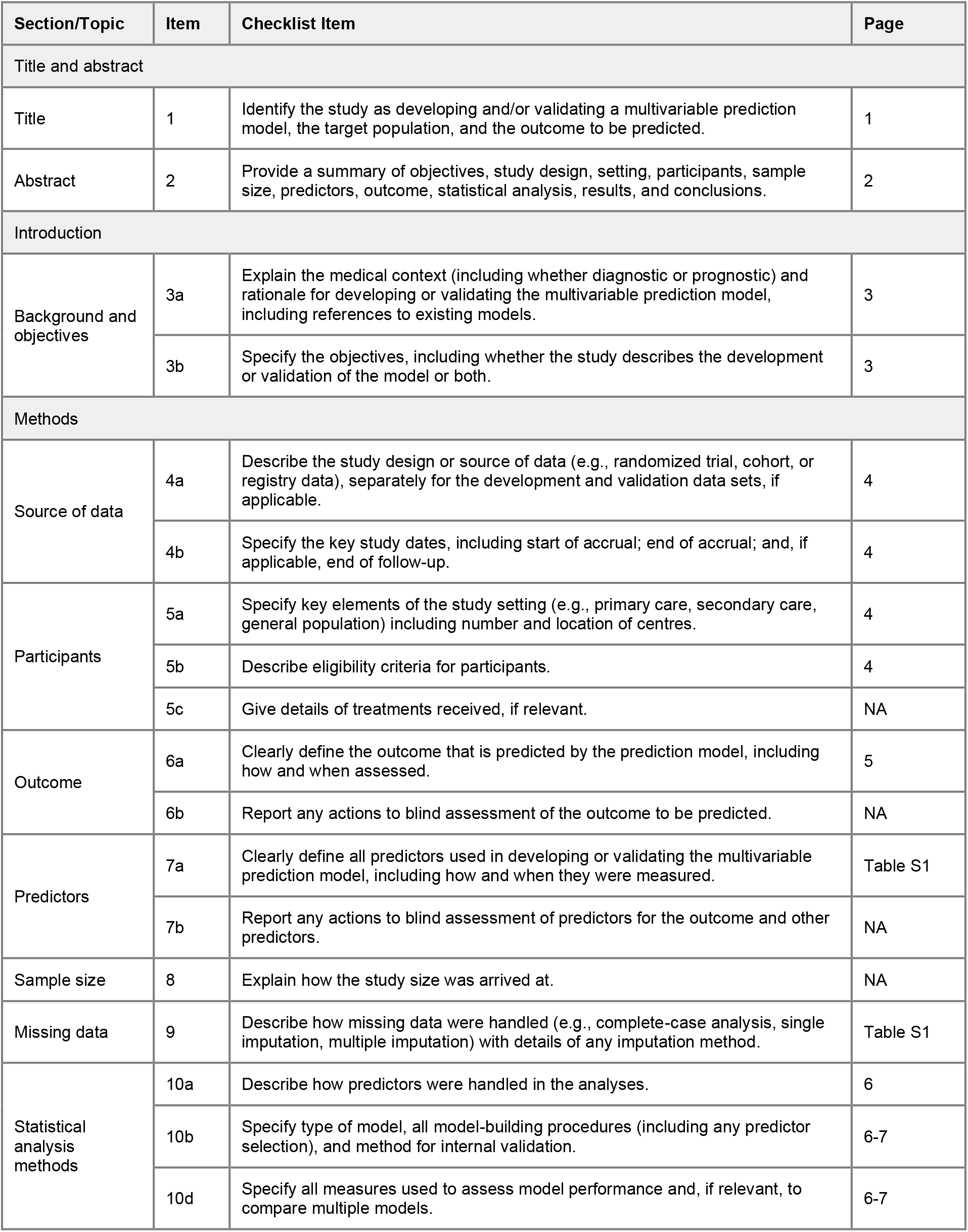
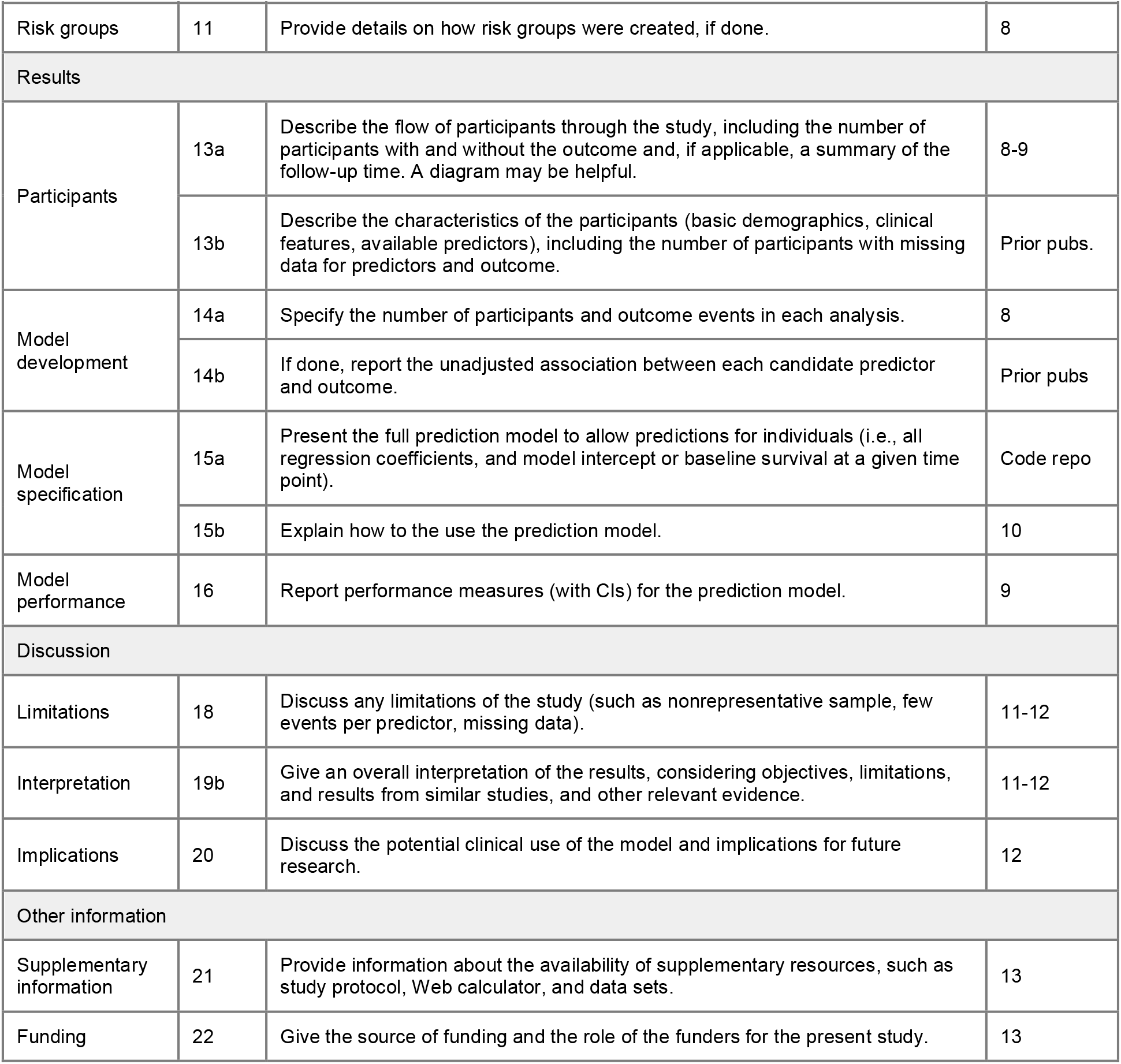
Transparent reporting of a multivariable prediction model for individual prognosis or diagnosis (TRIPOD) model reporting checklist.

| Section/Topic | Item | Checklist Item | Page |
| --- | --- | --- | --- |
| Title and abstract |  |  |  |
| Title | 1 | Identify the study as developing and/or validating a multivariable prediction model, the target population, and the outcome to be predicted. | 1 |
| Abstract | 2 | Provide a summary of objectives, study design, setting, participants, sample size, predictors, outcome, statistical analysis, results, and conclusions. | 2 |
| Introduction |  |  |  |
| Background and objectives | 3a | Explain the medical context (including whether diagnostic or prognostic) and rationale for developing or validating the multivariable prediction model, including references to existing models. | 3 |
|  | 3b | Specify the objectives, including whether the study describes the development or validation of the model or both. | 3 |
| Methods |  |  |  |
| Source of data | 4a | Describe the study design or source of data (e.g., randomized trial, cohort, or registry data), separately for the development and validation data sets, if applicable. | 4 |
|  | 4b | Specify the key study dates, including start of accrual; end of accrual; and, if applicable, end of follow-up. | 4 |
| Participants | 5a | Specify key elements of the study setting (e.g., primary care, secondary care, general population) including number and location of centres. | 4 |
|  | 5b | Describe eligibility criteria for participants. | 4 |
|  | 5c | Give details of treatments received, if relevant. | NA |
| Outcome | 6a | Clearly define the outcome that is predicted by the prediction model, including how and when assessed. | 5 |
|  | 6b | Report any actions to blind assessment of the outcome to be predicted. | NA |
| Predictors | 7a | Clearly define all predictors used in developing or validating the multivariable prediction model, including how and when they were measured. | Table S1 |
|  | 7b | Report any actions to blind assessment of predictors for the outcome and other predictors. | NA |
| Sample size | 8 | Explain how the study size was arrived at. | NA |
| Missing data | 9 | Describe how missing data were handled (e.g., complete-case analysis, single imputation, multiple imputation) with details of any imputation method. | Table S1 |
| Statistical analysis methods | 10a | Describe how predictors were handled in the analyses. | 6 |
|  | 10b | Specify type of model, all model-building procedures (including any predictor selection), and method for internal validation. | 6-7 |
|  | 10d | Specify all measures used to assess model performance and, if relevant, to compare multiple models. | 6-7 |
| Risk groups | 11 | Provide details on how risk groups were created, if done. | 8 |
| Results |  |  |  |
| Participants | 13a | Describe the flow of participants through the study, including the number of participants with and without the outcome and, if applicable, a summary of the follow-up time. A diagram may be helpful. | 8-9 |
|  | 13b | Describe the characteristics of the participants (basic demographics, clinical features, available predictors), including the number of participants with missing data for predictors and outcome. | Prior pubs. |
| Model development | 14a | Specify the number of participants and outcome events in each analysis. | 8 |
|  | 14b | If done, report the unadjusted association between each candidate predictor and outcome. | Prior pubs |
| Model specification | 15a | Present the full prediction model to allow predictions for individuals (i.e., all regression coefficients, and model intercept or baseline survival at a given time point). | Code repo |
|  | 15b | Explain how to use the prediction model. | 10 |
| Model performance | 16 | Report performance measures (with CIs) for the prediction model. | 9 |
| Discussion |  |  |  |
| Limitations | 18 | Discuss any limitations of the study (such as nonrepresentative sample, few events per predictor, missing data). | 11-12 |
| Interpretation | 19b | Give an overall interpretation of the results, considering objectives, limitations, and results from similar studies, and other relevant evidence. | 11-12 |
| Implications | 20 | Discuss the potential clinical use of the model and implications for future research. | 12 |
| Other information |  |  |  |
| Supplementary information | 21 | Provide information about the availability of supplementary resources, such as study protocol, Web calculator, and data sets. | 13 |
| Funding | 22 | Give the source of funding and the role of the funders for the present study. | 13 |

### SUPPLEMENTAL FIGURES

**Figure S1:**
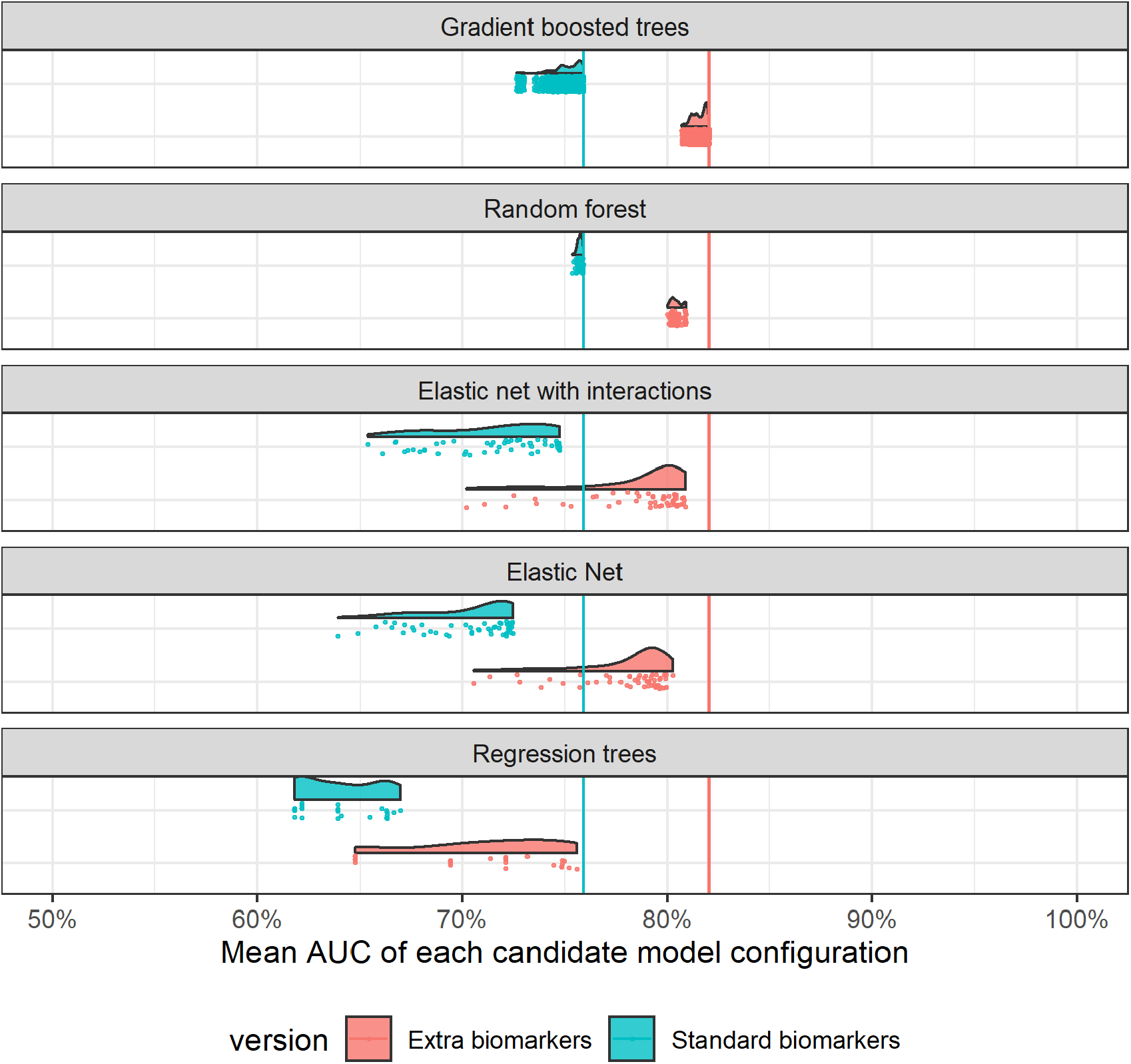
Average multiclass AUC for each evaluated model configuration as assessed using 5-fold cross validation and averaged across 15 tuning sets (excluding ensemble models). Each configuration (model type and hyperparameter set) is plotted as a dot. Distributions for each model type are shown.

**Figure S2:**
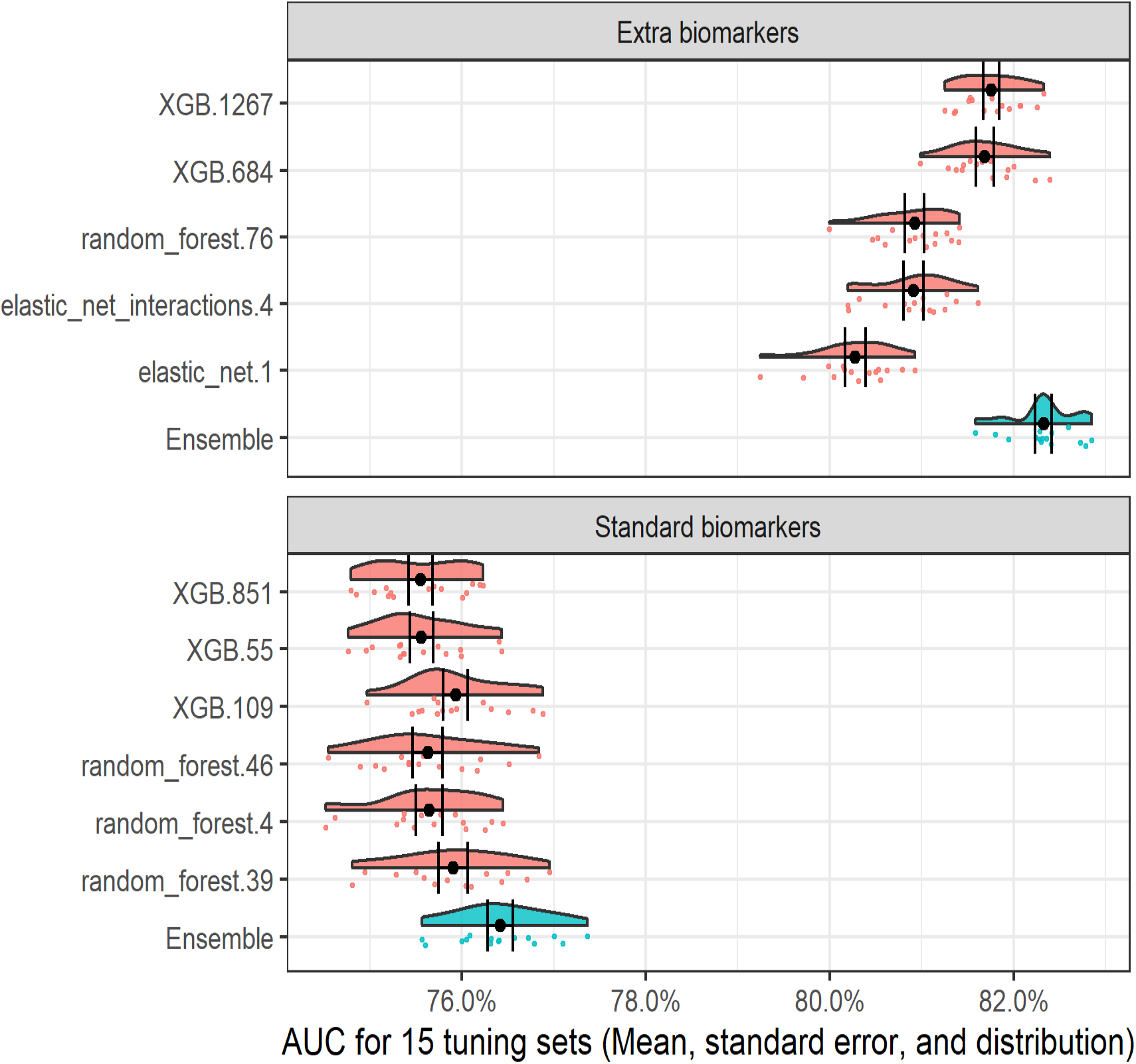
Distribution of multiclass AUC for across the 15 tuning sets for the top ensemble model configurations and the base model configurations that comprised them. For both the “standard” and “extra biomarkers” versions, the top ensemble was an average of the base models.

**Figure S3:**
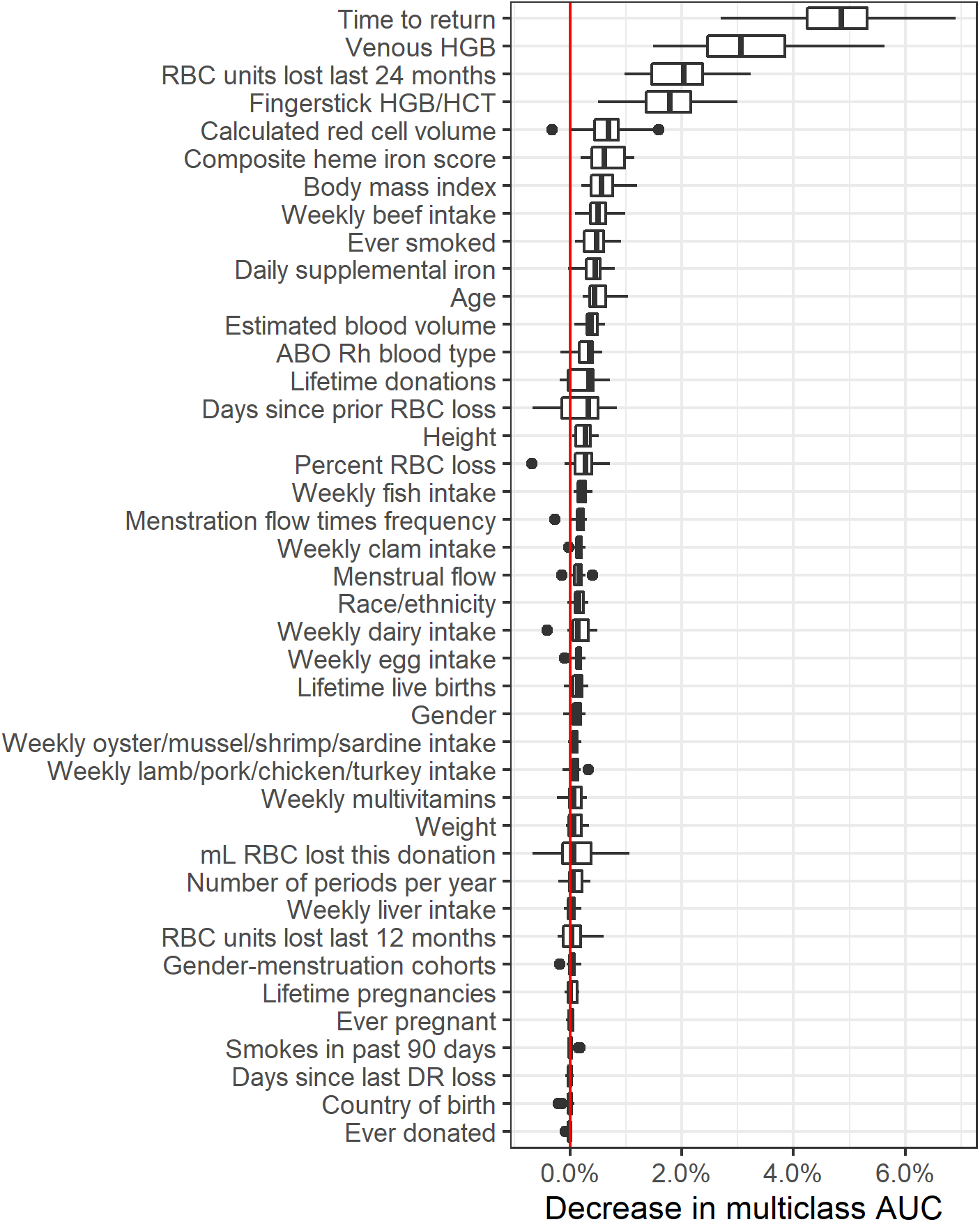
Relative variable importance for the top “standard biomarkers” model.

**Figure S4:**
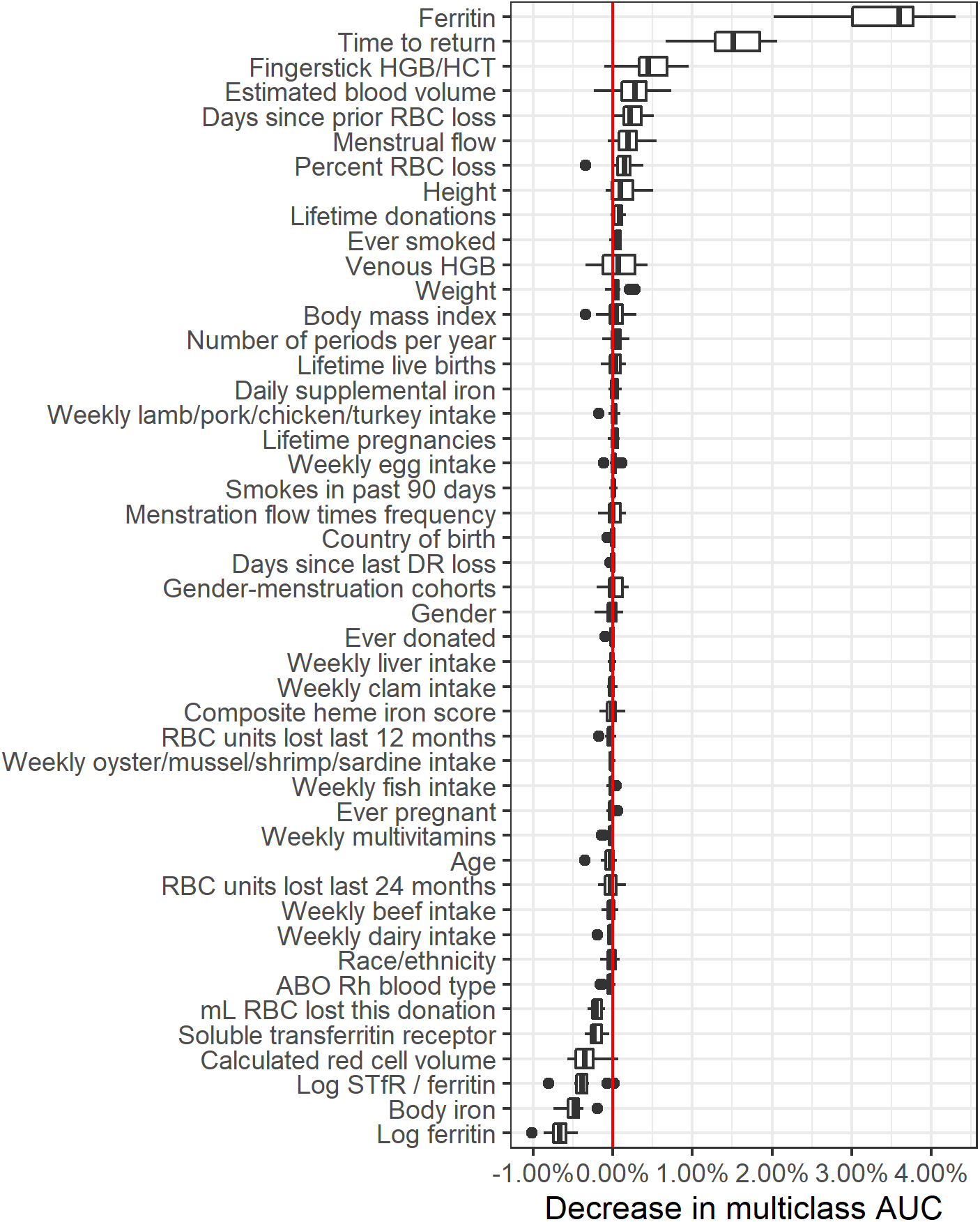
Relative variable importance for the top “extra biomarkers” model.

**Figure S5:**
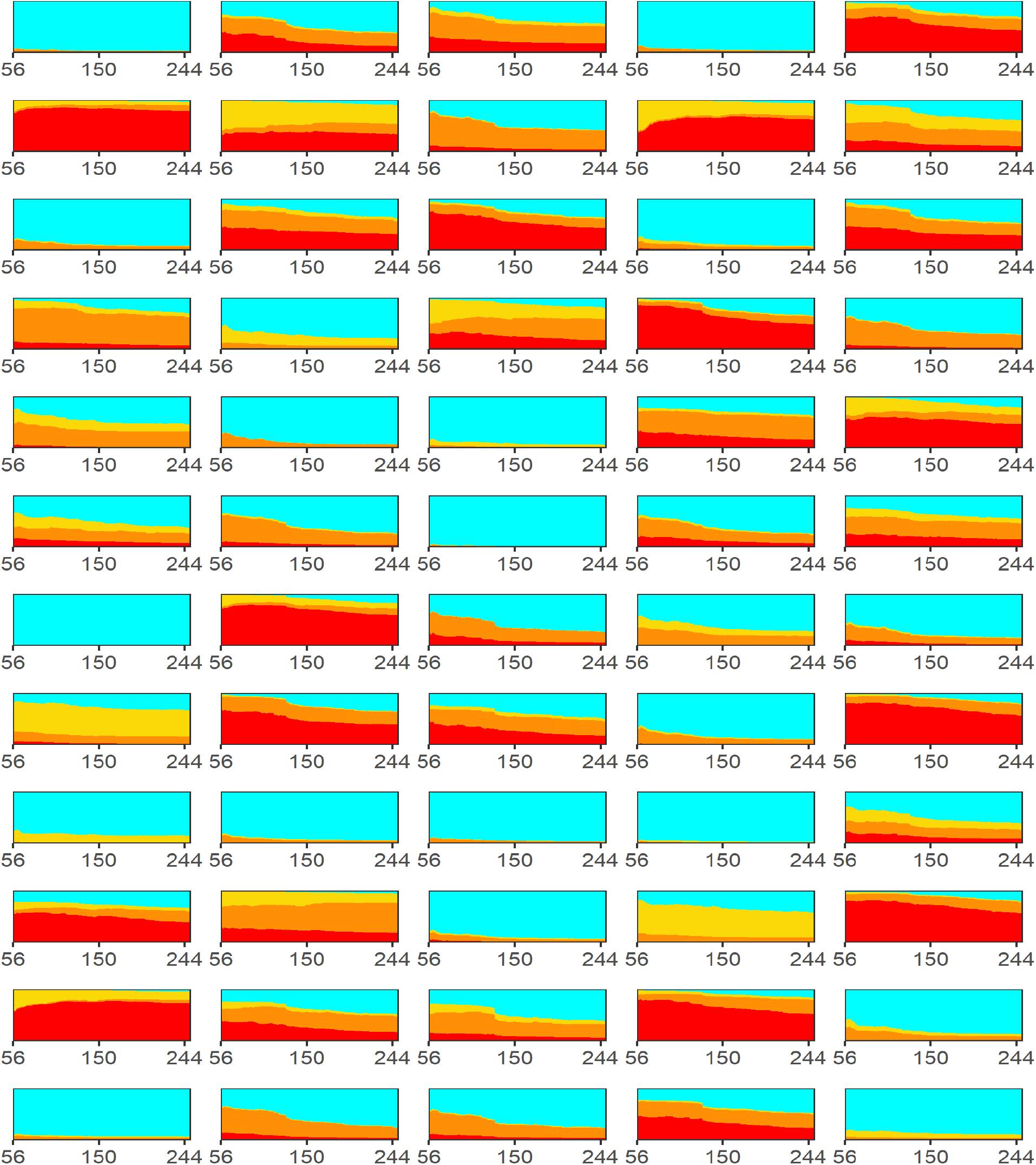
Individual risk trajectory for sixty randomly selected index donations. X-axis indicates the donation interval (days until a return donation attempt) and the height of the colored areas indicate the risk of each possible outcome: no adverse outcome (cyan), hemoglobin deferral (yellow), low iron donation (orange), and absent iron donation (red).

**Figure S6:**
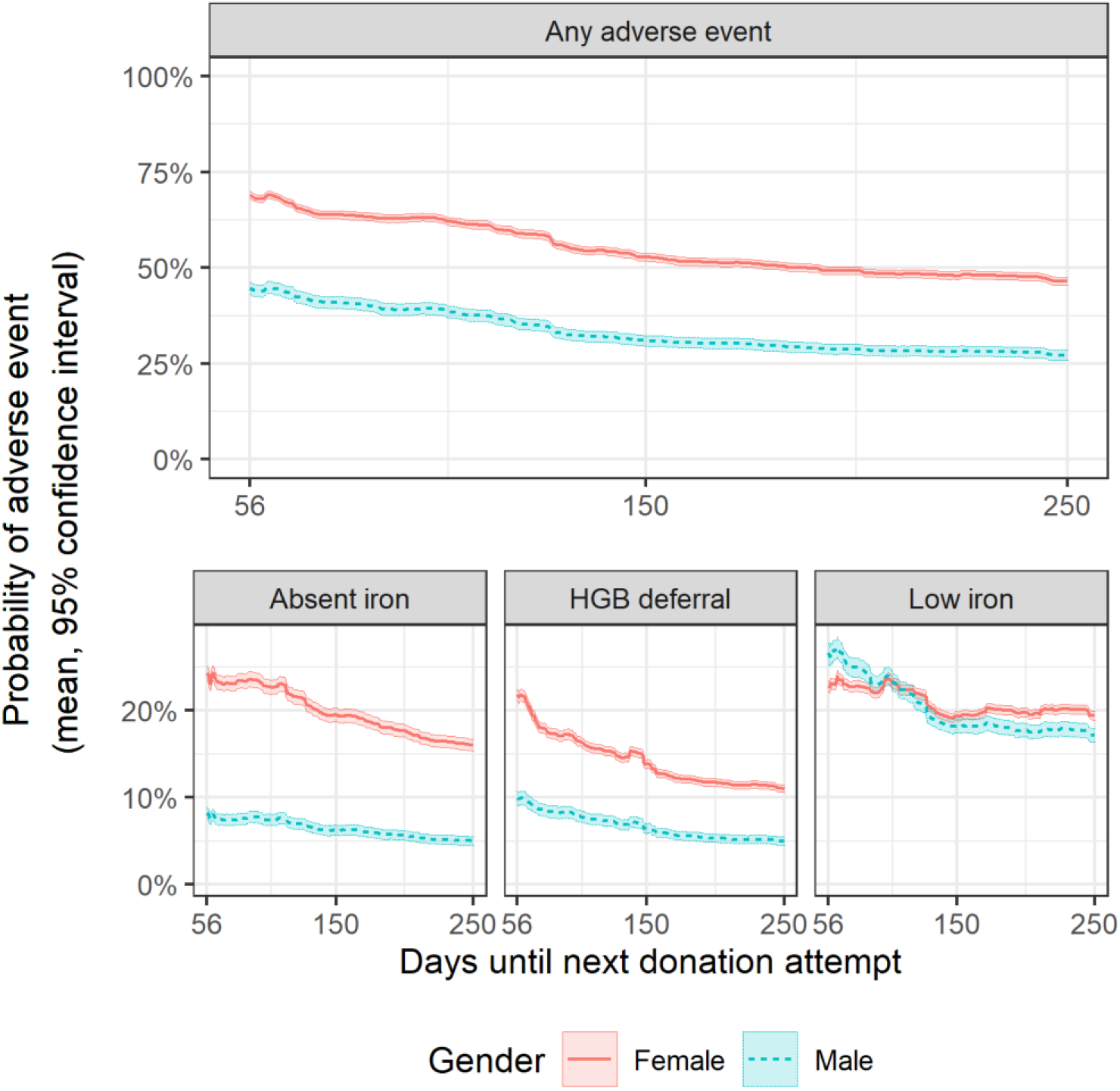
Average risk trajectory with 95% confidence intervals for donors in the first return dataset stratified by gender. Compared to men, women had higher estimated risk for absent iron donations and hemoglobin deferral but a similar average risk trajectory for low iron donations

**Figure S7:**
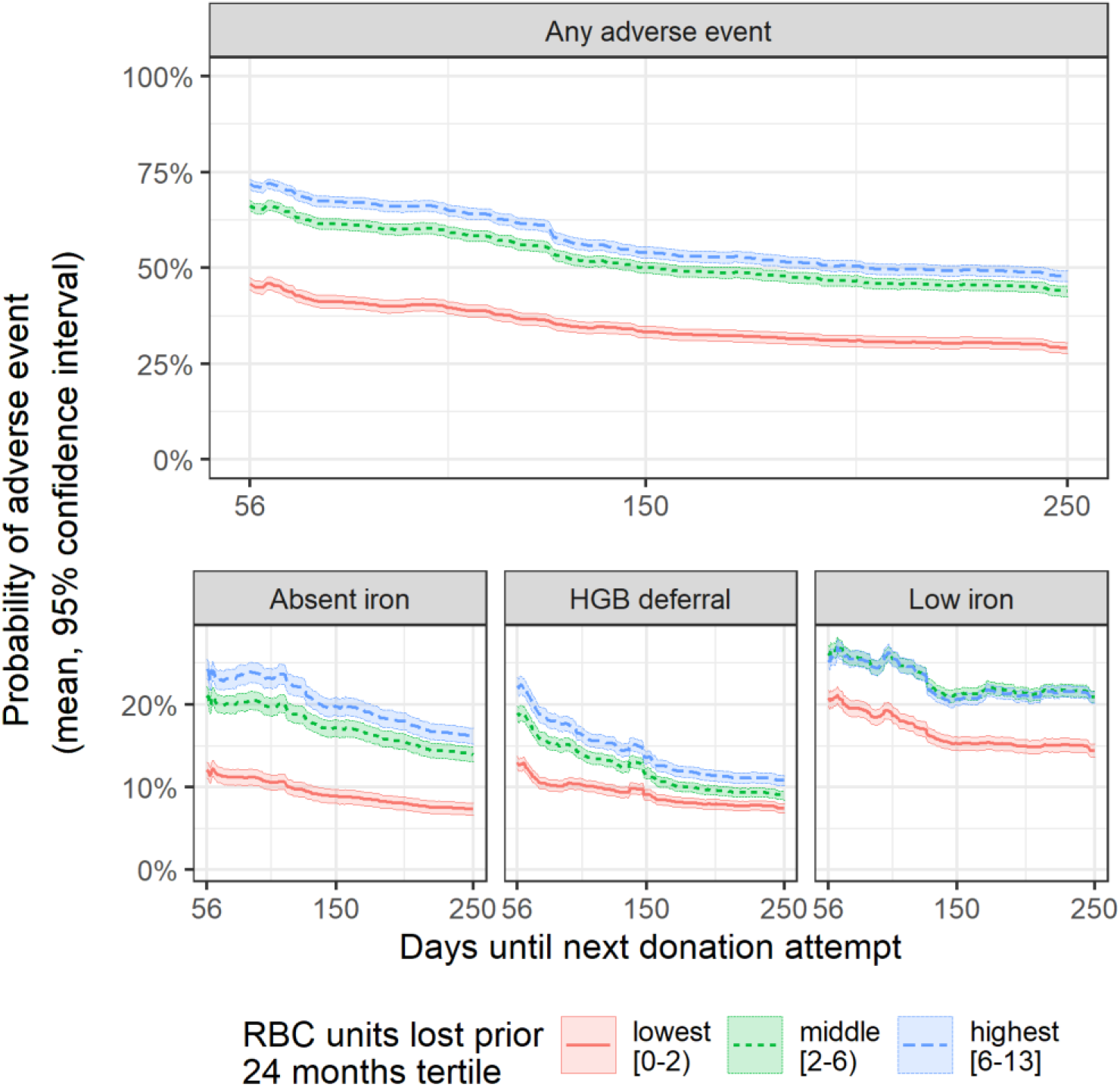
Average risk trajectory with 95% confidence intervals for donors in the first return dataset stratified by the number of red blook cell (RBC) units donated in the prior 24 months. Those who donated 2 or fewer units in the prior two years had lower risk of adverse outcomes, particularly absent iron donations.

**Figure S8:**
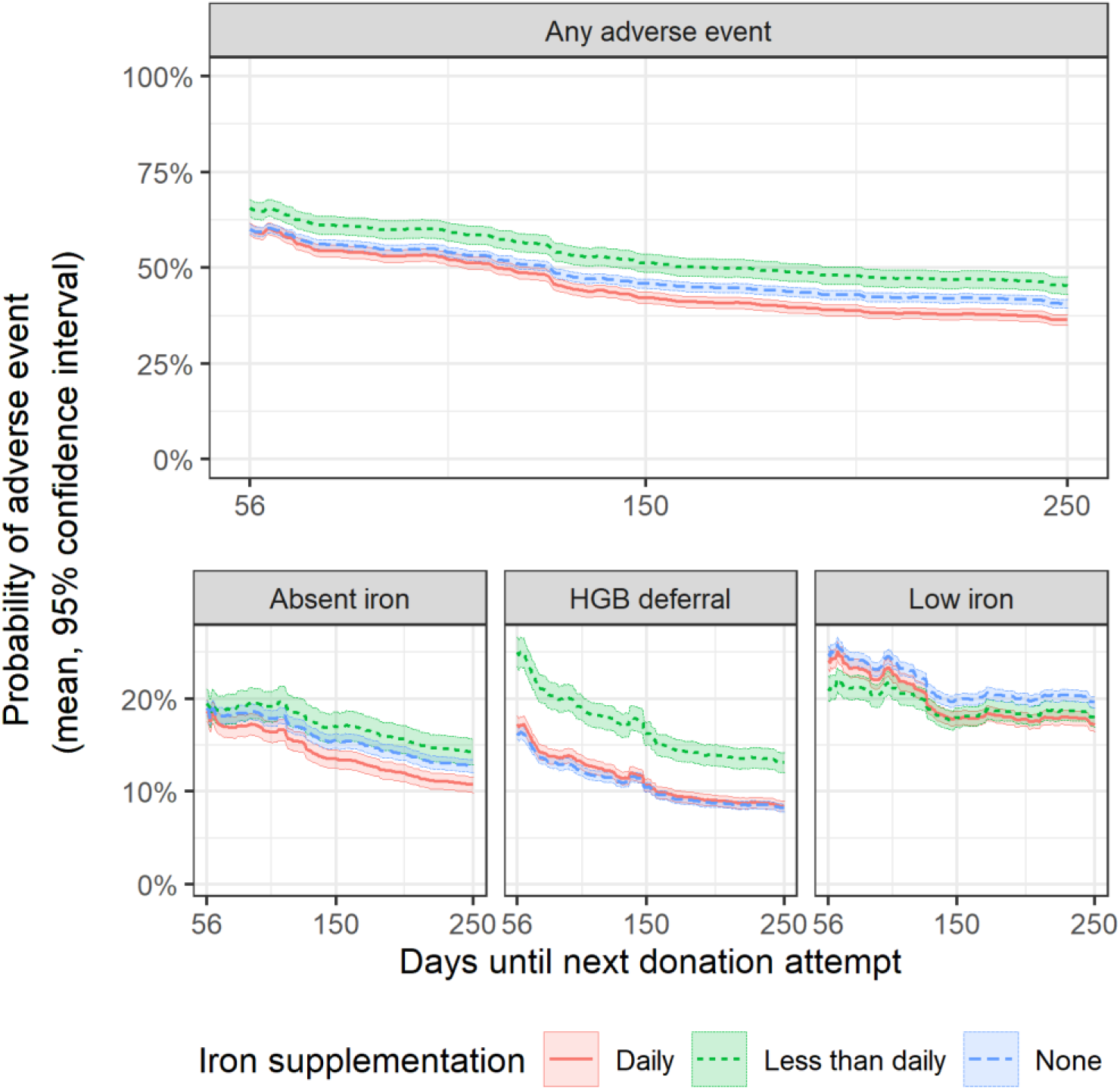
Average risk trajectory with 95% confidence intervals for donors in the first return dataset stratified by iron supplementation. Donors with ‘less than daily’ iron supplementation had lower risk of adverse outcomes, particularly hemoglobin deferral, whereas donors taking either no iron supplmeentation or daily iron supplementation had more similar risk trajectories. These results are not intuitive, but may be due to confounding variables, for which this analysis does not account. For example, donors with diagnosed anemia or a related condition may be more likely to take daily iron supplementation.

**Figure S9:**
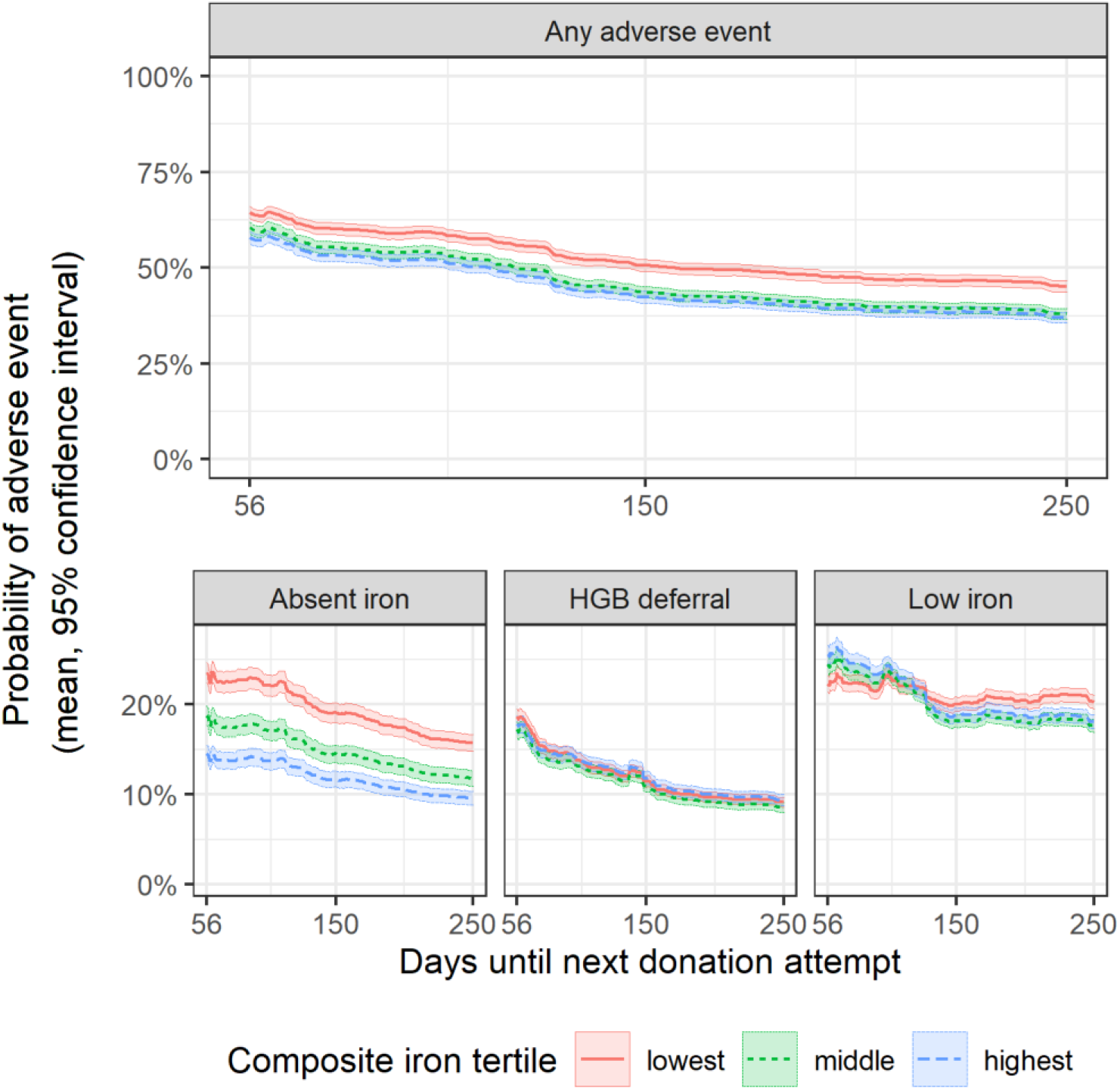
Average risk trajectory with 95% confidence intervals for donors in the first return dataset stratified by heme dietary iron intake score, which is calculated from self-reported dietary data, at index donation. On average, donors in the lowest tertile of heme iron intake had a higher estimated risk of an absent iron donation but similar risk trajectory for hemoglobin deferral or a low iron donation.

